# Molecular subgroups of ALS patients with distinct survival outcomes identified through plasma proteomics

**DOI:** 10.64898/2026.09.16.26362998

**Authors:** Inci S. Aksoylu, Louisa Azizi, Linn Öijerstedt, Sofia Lilja, María Bueno Álvez, Solmaz Yazdani, Lovisa Skoglund, Juliette Foucher, Alexander Juto, Christina Seitz, Rayomand Press, Kristin Samuelsson, Ulf Kläppe, Fang Fang, Mathias Uhlén, Fredrik Edfors, Peter Nilsson, Caroline Ingre, Anna Månberg

**Affiliations:** Department of Protein Science, KTH Royal Institute of Technology, SciLifeLab, Stockholm, Sweden; Department of Clinical Neuroscience, Karolinska Institutet, Stockholm, Sweden; Department of Neurology, Karolinska University Hospital, Stockholm, Sweden; Institute of Environmental Medicine, Karolinska Institutet, Stockholm, Sweden

## Abstract

Amyotrophic Lateral Sclerosis (ALS) is a phenotypically diverse neurodegenerative disorder characterized by the degeneration of motor neurons, ultimately resulting in loss of motor function. Currently, patients are classified based on clinical factors which does not fully capture the heterogeneity. Using the Olink Explore HT, we profiled ∼5400 plasma proteins in ALS patients (n = 235) from the ALSrisc Study at Karolinska Institutet, Sweden, and applied consensus clustering, which revealed three patient subgroups significantly differing in survival outcome. Weighted gene correlation network analysis (WGCNA) and gene ontology (GO) analysis identified protein modules related to biological functions including cellular stress response and Rho-GTPase activity for the shorter surviving patient group. Cell-type enrichment analysis indicating a broad cellular origin of the circulating proteome, with contributions from nervous system cells, vascular cells, immune cells, and muscle cells. Our work highlights the potential of plasma proteomics for clinically meaningful patient stratification in ALS.

## MAIN TEXT

Amyotrophic Lateral Sclerosis (ALS) is a heterogenous neurodegenerative disorder which affects upper and lower motor neurons leading to the loss of motor function, clinically manifesting as progressive muscle weakness, leading to paralysis and respiratory failure^1,2^. ALS-patients are still being diagnosed and classified through clinical examination, assessing site of symptom onset, degree of the upper and lower motor neuron involvement in combination with degree of motor function^1,3^. Growing evidence suggests ALS to be a far more biologically and phenotypically complex disorder than can be explained relying only on clinical information overlooking the underlying molecular heterogeneity^4,5^. Improved knowledge of a more biology-driven, molecular patient classification could gain insights enhancing patients’ stratification, important also in the development of targeted treatments for ALS.

Recent advances in large-scale platforms such as Olink Explore HT,SomaScan and NULISA have expanded the use of proteomics in clinical research by enabling multiplex measurement of both high- and low-abundance proteins across large patient cohorts^6–11^. Several studies using these platforms to profile the plasma proteome in ALS have uncovered alterations in the abundances of proteins involved in diverse biological processes, including muscle development, cellular homeostasis, and neuronal function^9,12–14^. Changes in plasma profiles have also been identified in early cases, supporting the use of plasma proteomics in detection of presymptomatic ALS^15^. Even though these studies offer valuable insight into plasma proteome changes, they mainly rely on case-control comparisons and therefore do not capture the underlying ALS-heterogeneity. Previous studies aiming to characterize molecular heterogeneity have largely depended on post-mortem tissue, limiting their relevance for living patients and for tracking disease progression over time^5,16–21^. Altogether, the urgent need for minimally invasive biomarkers that capture molecular heterogeneity and have relevance both for prognosis and future clinical trials remains to be addressed.

The Swedish case-control ALSrisc Study was initiated in 2016 at the ALS Clinical Research Centre at Karolinska Institutet, Stockholm, with the aim to improve understanding of ALS risk and progression as well as identifying disease-related biomarkers^22^. In the presented work, we used the Olink Explore HT platform for relative quantification of ∼5400 proteins in combination with consensus clustering to identify plasma prote-omics-based subgroups among patients enrolled in the ALSrisc Study. The observed groups were found to differ in survival outcomes, and their associated protein modules were mapped to biological processes that include but are not limited to cellular stress and metabolism. By annotating proteins according to their cell-type enriched expression, we demonstrate that the subgroup-associated plasma profiles reflect contributions from various cell-types beyond those of central nervous system origin. Collectively, our findings present a novel, biology-based patient stratification framework that links plasma proteome alterations to disease biology and differences in patient survival.

## Results

In this study, we used large scale plasma proteomics data of 235 patient samples to identify and characterize molecular patient subgroups (**Table 1**). Out of the ∼5400 proteins analyzed, 2349 were detected above the limit of detection (LOD) in at least 85% of the samples and remained after quality control and were retained for subsequent analysis.

**Table 1:** Characteristics of the study cohort. Demographic and clinical characteristics (age, first visit ALSFRS-R score, survival duration in months, disease progression category and site of onset) presented for 235 patients stratified by sex (n_Female_= 103, n_Male_ = 132). Continuous variables are reported as mean (min-max) and categorical variables as n (%). No statistically significant differences were observed between sexes for any of the variables. P-values were calculated using Wilcoxon rank sum test, Pearson’s Chi-squared test, or Fisher’s exact test as appropriate.

| VARIABLE | OVERALL<br>N = 235<sup>1</sup> | SEX |  | P-VALUE<sup>2</sup> |
| --- | --- | --- | --- | --- |
|  |  | FEMALE<sup>†</sup> | MALE<sup>†</sup> |  |
| NA |  |  |  |  |
| Age | 66 (24–97) | 67 (37–97) | 64 (24–84) | 0.087 |
| ALSFRS-R Score (First visit) | 38 (18–48) | 37 (18–46) | 39 (18–48) | 0.082 |
| Survival (Months) | 36 (3–200) | 37 (6–103) | 36 (3–200) | 0.467 |
| PROGRESSION |  |  |  |  |
| Fast | 75 (32%) | 27 (26%) | 48 (36%) | 0.098 |
| Slow | 38 (16%) | 18 (17%) | 20 (15%) | 0.631 |
| Intermediate | 122 (52%) | 58 (56%) | 64 (48%) | 0.234 |
| ONSET |  |  |  |  |
| Bulbar | 87 (37%) | 45 (44%) | 42 (32%) | 0.061 |
| Spinal | 135 (57%) | 54 (52%) | 81 (61%) | 0.169 |
| FTD | 5 (2.1%) | 2 (1.9%) | 3 (2.3%) | >0.999 |
| Other | 8 (3.4%) | 2 (1.9%) | 6 (4.5%) | 0.471 |
| <sup>†</sup> Mean (Min–Max); n (%) |  |  |  |  |
| <sup>2</sup> Wilcoxon rank sum test; Pearson's Chi-squared test; Fisher's exact test |  |  |  |  |

### Consensus clustering of the plasma proteome profiles reveals three subgroups of ALS patients

Consensus clustering of patients was performed using normalized protein expression (NPX) values adjusted for age, sex and median NPX per patient as these covariates were identified as significantly associated with global protein patterns (**Suppl. Fig. 1**). Using the 75% most variable proteins (n = 1762) allowed us to identify the optimal clustering strategy across multiple combinations of feature-selection and clustering methods, as well as optimal number of clusters. Our analysis revealed three stable subgroups of ALS patients, referred to as Class 1-3, with significantly different plasma profiles across 1466 out of the 1762 proteins (FDR < 0.05, |Log_2_Fold-change| > 0.25) (**Suppl. Fig. 2**). Class 1 and 2 patients exhibited more homogeneous abundance patterns, whereas Class 3 patients showed greater heterogeneity across the 1466 proteins compared to the other two subgroups, showing more extreme profiles (**Fig. 1A**). Visualizing the data using Uniform Manifold Approximation and Projection (UMAP) to project adjusted NPX values across proteins into a low-dimensional space also illustrated a separation of Class 2 (n = 66) from Class 1 (n = 68) and Class 3 (n=101) patients (**Fig. 1B**).

**Figure 1:**
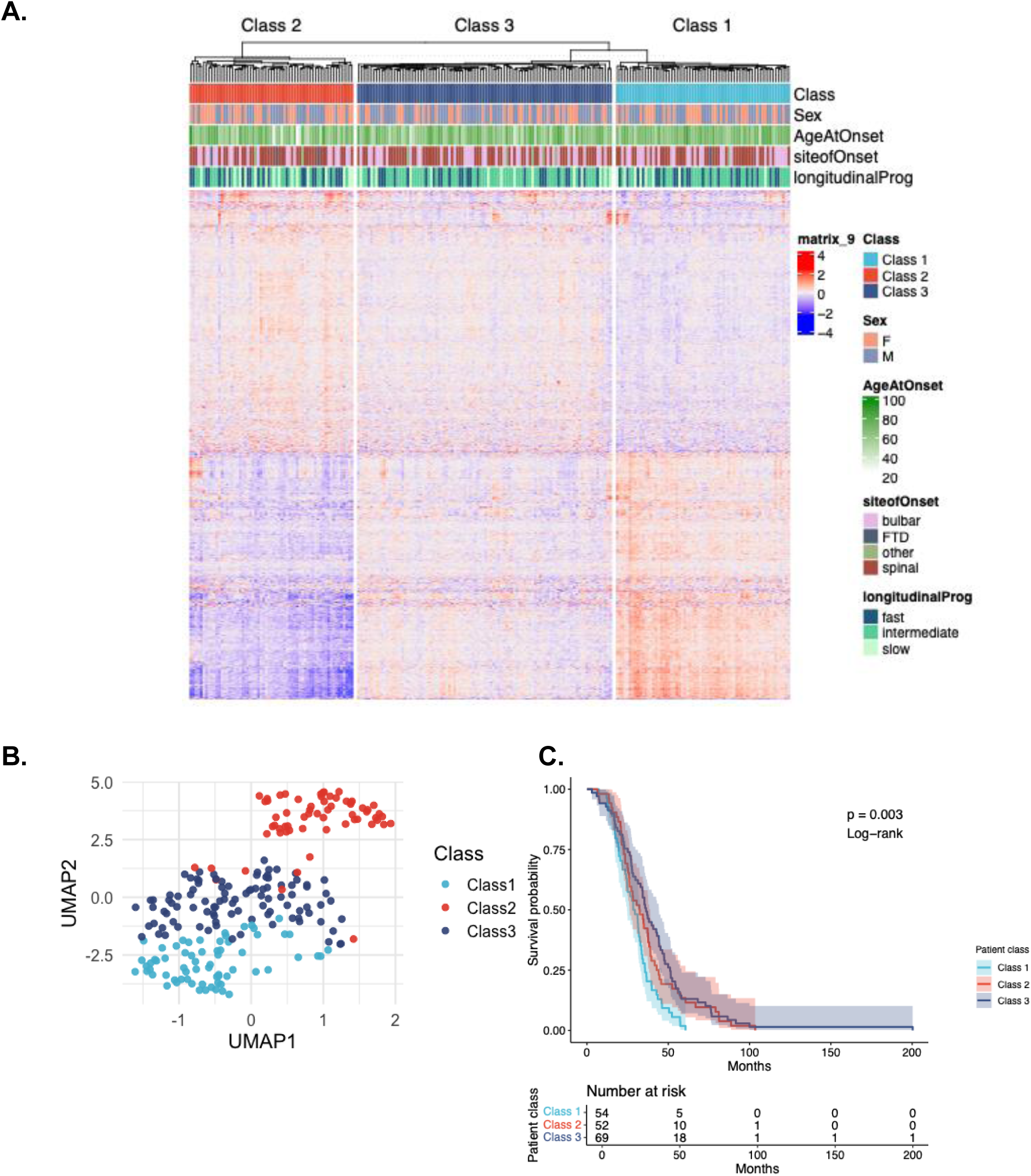
Unsupervised clustering reveals subgroups of ALS. **a.** Heatmap showing unsupervised clustering of ALS patients (n=228) based on plasma proteome profiles where columns represent patients and rows represent all proteins that significantly (FDR < 0.05, |log2FC| > 0.25, n = 1466) differ between patient subgroups. Heatmap values represent residual abundance of proteins and red indicates high abundance while blue indicates low abundance. Annotation bars above the heatmap from top to bottom indicate patient subgroup (Class), sex, age at symptom onset (”ageatOnset”), site. of symptom onset (“siteofOnset”) and disease progression category (“longitudinalProg”). **b.** Uniform Manifold Approximation and Projection (UMAP) visualization of patient subgroups generated using significantly altered proteins where each colored dot represents an individual while colors encode different patient subgroups. **c.** Kaplan-Meier survival curves for identified ALS subgroups. Error bands shaded differently for each subgroup represents 95% confidence intervals, p-values were calculated using Log-rank test and the table below represents the number of individuals at risk for each patient subgroup.

We further assessed clinical characteristics of subgroups including age of onset, site of symptom onset, disease progression rate and survival (**Table 2**). We observed that age, site of onset and sex were not significantly different across patient subgroups, although Class 1 patients had a marginally higher median age of onset when compared Class 2 (p = 0.06, Wilcoxon rank sum test, **Suppl. Fig 3A**) and Class 3 (p = 0.14, Wilcoxon rank sum test, **Suppl. Fig. 3A**) (Class 1: 68 years, Class 2: 64, Class 3: 65 years, p = 0.161, Kruskal-Wallis Test, **Suppl. Fig. 3A**). Class 3 had a higher proportion of patients with intermediate progression rate when compared to the other subgroups (Class 1: 52%, Class 2: 38%, Class 3: 59%, p = 0.032, Pearson’s Chi-squared test), while Class 2 had a higher proportion of slow progressing patients (Class 1: 12%, Class 2: 25%, Class 3: 14%, p = 0.105, Pearson’s Chi-squared test). Furthermore, we observed significant differences in mean survival time across the three subgroups with patients in Class 1 having the lowest mean survival time (Class 1: 30 months, Class 2: 37 months, Class 3: 41 months, p = 0.027, Kruskal-Wallis test, **Table 2**). On the other hand, site of onset did not differ significantly across patient subgroups, except for FTD onset, which was significantly different (p = 0.037,Fisher’s exact test, **Table 2**) but represented only a small number of patients within the cohort (n = 4, 1.8%). We then performed Kaplan-Meier survival analysis to assess survival outcomes of identified patient subgroups. Our analysis confirmed significant differences in survival outcomes with patients in Class 1 exhibiting the poorest survival (p = 0.003, Log-rank test) (**Fig. 1C**). Notably, patients in Class 1 had shorter survival than Class 2 and Class 3 patients despite having a higher mean first-visit ALSFRS-R score than Class 2 patients (p < 0.01, Wilcoxon rank sum test, **Table 2**) and a similar score to Class 3 patients (p > 0.05, Wilcoxon rank sum test; **Table 2, Suppl. Fig. 3B**). Furthermore, NEFL, an established circulating prognostic marker of ALS, was significantly elevated in Class 2 patients compared to Class 1 (p = 0.0085, Wilcoxon rank sum test, **Suppl. Fig. 3C**), with no significant difference observed between Class 2 and Class 3 (p = 0.059, Wilcoxon rank sum test) (p = 0.027, Kruskal-Wallis Test; **Suppl. Fig. 3C**)^23^.

**Table 2:** Characteristics of the study cohort stratified by patient subgroups. Demographic and clinical characteristics of 228 confidently-clustered (silhouette score > 0.7) patients distributed across three subgroups. Continuous variables are reported as mean (min-max) and categorical variables as n (%). P-values were calculated using Wilcoxon rank sum test, Pearson’s Chi-squared test, or Fisher’s exact test as appropriate. Statistically significant differences in ALSFRS-R score at first visit (p = 0.014), survival duration (p = 0.027) and progression type (p = 0.032) were observed across patient subgroups.

| VARIABLE | OVERALL<br>N = 228<sup>1</sup> | PATIENT CLASS |  |  | P-VALUE<sup>2</sup> |
| --- | --- | --- | --- | --- | --- |
|  |  | CLASS 1<sup>1</sup> | CLASS 2<sup>1</sup> | CLASS 3<sup>1</sup> |  |
| NA |  |  |  |  |  |
| Age | 66 (24–97) | 68 (45–87) | 64 (24–97) | 65 (37–85) | 0.161 |
| ALSFRS-R Score (First visit) | 38 (18–48) | 39 (18–48) | 36 (18–47) | 39 (22–47) | **0.014** |
| Survival (Months) | 36 (3–200) | 30 (7–61) | 37 (6–103) | 41 (3–200) | **0.027** |
| SEX |  |  |  |  |  |
| Female | 100 (44%) | 31 (46%) | 32 (51%) | 37 (38%) | 0.238 |
| Male | 128 (56%) | 36 (54%) | 31 (49%) | 61 (62%) | 0.238 |
| PROGRESSION |  |  |  |  |  |
| Fast | 73 (32%) | 24 (36%) | 23 (37%) | 26 (27%) | 0.304 |
| Slow | 38 (17%) | 8 (12%) | 16 (25%) | 14 (14%) | 0.085 |
| Intermediate | 117 (51%) | 35 (52%) | 24 (38%) | 58 (59%) | **0.032** |
| ONSET |  |  |  |  |  |
| Bulbar | 84 (37%) | 27 (40%) | 21 (33%) | 36 (37%) | 0.713 |
| Spinal | 132 (58%) | 38 (57%) | 37 (59%) | 57 (58%) | 0.971 |
| FTD | 4 (1.8%) | 1 (1.5%) | 3 (4.8%) | 0 (0%) | **0.037** |
| Other | 8 (3.5%) | 1 (1.5%) | 2 (3.2%) | 5 (5.1%) | 0.486 |

<sup>1</sup> Mean (Min–Max); n (%)
<sup>2</sup> Kruskal-Wallis rank sum test; Pearson’s Chi-squared test; Fisher’s exact test

### Weighted gene correlation network analysis identifies plasma protein modules associated with patient subgroups

We applied weighted gene correlation network analysis (WGCNA) to the 1466 proteins that differed across patient subgroups to identify protein correlation modules and assess their association with patient subgroups. Using a soft-thresholding power of 4 followed by hierarchical clustering, we identified nine modules (n_Turquoise_= 566, n_Blue_= 268, n_Yellow_=53, n_Black_=42, n_Brown_=102, n_Green_= 44, n_Pink_=25, n_Red_= 44), including a grey module (n_grey_=322) comprising proteins that could not be assigned to any other module due to low connectivity (**Suppl. Fig. 4, Suppl. File 1**). Module eigengenes (MEs) values representing the first principal component of each module, were correlated to patient subgroups. Modules that positively correlate with Class 1 patients included the green, red, turquoise modules, while Class 2 patients showed positive correlations with the black, blue, pink and brown modules (**Fig. 2A-B, Suppl. Fig. 5A**). Consistent patterns were also observed for hub proteins, such as ASCC1 in turquoise module or TNFRSF9 in brown module (**Suppl. Fig. 5C-D**).

**Figure 2:**
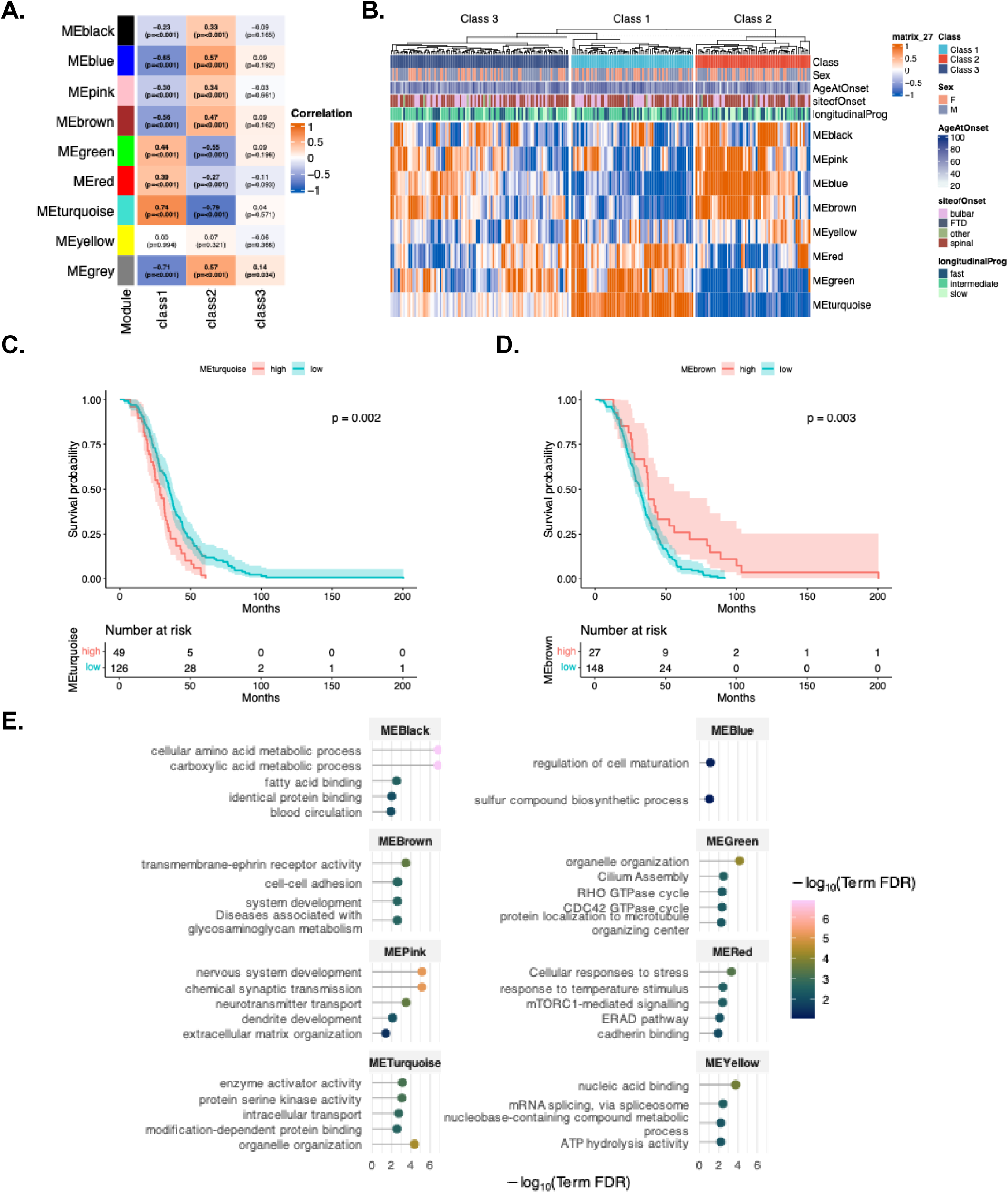
Weighted Gene Correlation Network Analysis (WGCNA) modules relate to patient subgroups and survival. **a.** Heatmap showing module-trait association between identified protein modules (n_Turquoise_= 566, n_Blue_= 268, n_Yellow_=53, n_Black_=42, n_Brown_=102, n_Green_= 44, n_Pink_=25, n_Red_= 44) and patient subgroups where rows represent co-expression modules and columns represent patient subgroups. Coloring of the heatmap corresponds to the strength and direction of association between the module eigengenes (MEs) and the traits. Values within cells represent the Pearson correlation coefficient (r) and Student asymptotic p-values are shown in parentheses and statistically significant (p<0.05) correlations are indicated in bold. **b.** Heatmap showing module abundances across patients and patient subgroups. Heatmap values represent per patient ME values, red indicates high abundance while blue indicates low abundance.Annotation bars above the heatmap from top to bottom indicate patient subgroup (Class), sex, age at symptom onset (”ageatOnset”), site of symptom onset (“siteofOnset”) and disease progression category(“longitudinalProg”). **c.** and **d.** Kaplan-Meier survival curves stratified into “high” (in red) and “low” (in teal) groups by turquoise and blue ME values, respectively. The p-values were calculated using Log-rank test, error bands represent 95% confidence interval and tables below represent number of individuals at risk for “high” and “low” groups. **e.** Dot plots showing five most significant terms (GO-BP, GO-MF and Reactome Pathways) from functional enrichment analysis for proteins within each WGCNA module. Each panel corresponds to a module. Coloring of dots and the x-axis represent −log_10_(FDR) value where higher values reflect stronger enrichment.

To evaluate the prognostic relevance of these modules, patients were stratified into “high” and “low” abundance groups based on optimal ME cutoffs, and survival was assessed using Kaplan-Meier analysis. Higher levels of turquoise, red and green modules that positively correlated with Class 1 were linked to poorer survival (p = 0.002, p = 0.011 and p=0.040, Log-rank test, respectively; **Suppl. Fig. 5B**), whereas higher levels of blue, brown and pink modules that positively correlated with Class 2 were associated with longer survival, except for the black module (p = 0.008, p = 0.003, p = 0.004, Log-rank test, respectively; **Suppl. Fig. 5B**). The strongest associations to survival outcome were observed for the turquoise and brown modules (p=0.002 and 0.003, Log-rank test, respectively) (**Fig. 2C-D**).

To link protein modules to biological functions, we performed functional enrichment analysis using Gene Ontology (GO) and REACTOME pathways. We observed that the modules associated with shorter survival were enriched for processes including Rho and CDC42 GTPase cycle (green, **Suppl. Fig. 6E**), cellular response to stress (red, **Suppl. Fig. 6F**) and protein-serine kinase activity (turquoise, **Suppl. Fig. 6G**). In contrast, modules associated with longer survival were enriched with terms such as acid metabolism and circulation (black, **Suppl. Fig. 6A**), synaptic transmission (pink, **Suppl. Fig. 6C**) and cell maturation (blue, **Suppl. Fig. 6B**) (**Fig. 2E**, **Suppl. File 1**). Notably, the yellow module enriched for RNA metabolism, a process increasingly seen as central to ALS, showed no significant association with any patient subgroup yet remained predictive of survival (**Suppl. Fig. 5B**)^24–26^.

### Cell-type related markers differentiate between subgroups of ALS

By leveraging transcriptomics data from the Human Protein Atlas single cell resource, we identified proteins enriched in 154 cell types. Following this, we assessed whether individual cell types contribute to subgroup specific plasma profiles using type II analysis of variance (ANOVA), with effect sizes estimated by partial eta squared (η^2^), reflecting the proportion of variance in each marker explained by the subgroups. We observed that the plasma levels of 30 cell-type enriched markers differed significantly across patient subgroups, suggesting that multiple cell types might contribute to the altered plasma proteome (**Fig. 3A, Suppl. Fig 7, Suppl. Fig 8A, Suppl. File 2**), with the most significant being GRAP2 (platelets) for Class 1 and DLL1 (ependymal cells) for Class 2 patients (**Fig. 3B, Suppl. Fig. 7, Suppl. Fig. 8A-B**). Markers with increased abundance in Class 1 patients suggested contributions from brain-resident and adaptive immune cell populations, including excitatory neurons, oligodendrocytes and T-cells. Class 2 patients, in contrast, showed greater contributions from innate immune cells such as mast cells and monocytes, as well as cells related to vascular function such as vascular endothelial cells and pericytes (**Suppl. Fig. 7, Suppl. Fig. 8B**). While Class 1 and Class 2 patients showed opposing abundance patterns for the identified cell-type related markers, Class 3 patients rather displayed heterogenous patterns across these markers (**Fig. 3C**).

**Figure 3:**
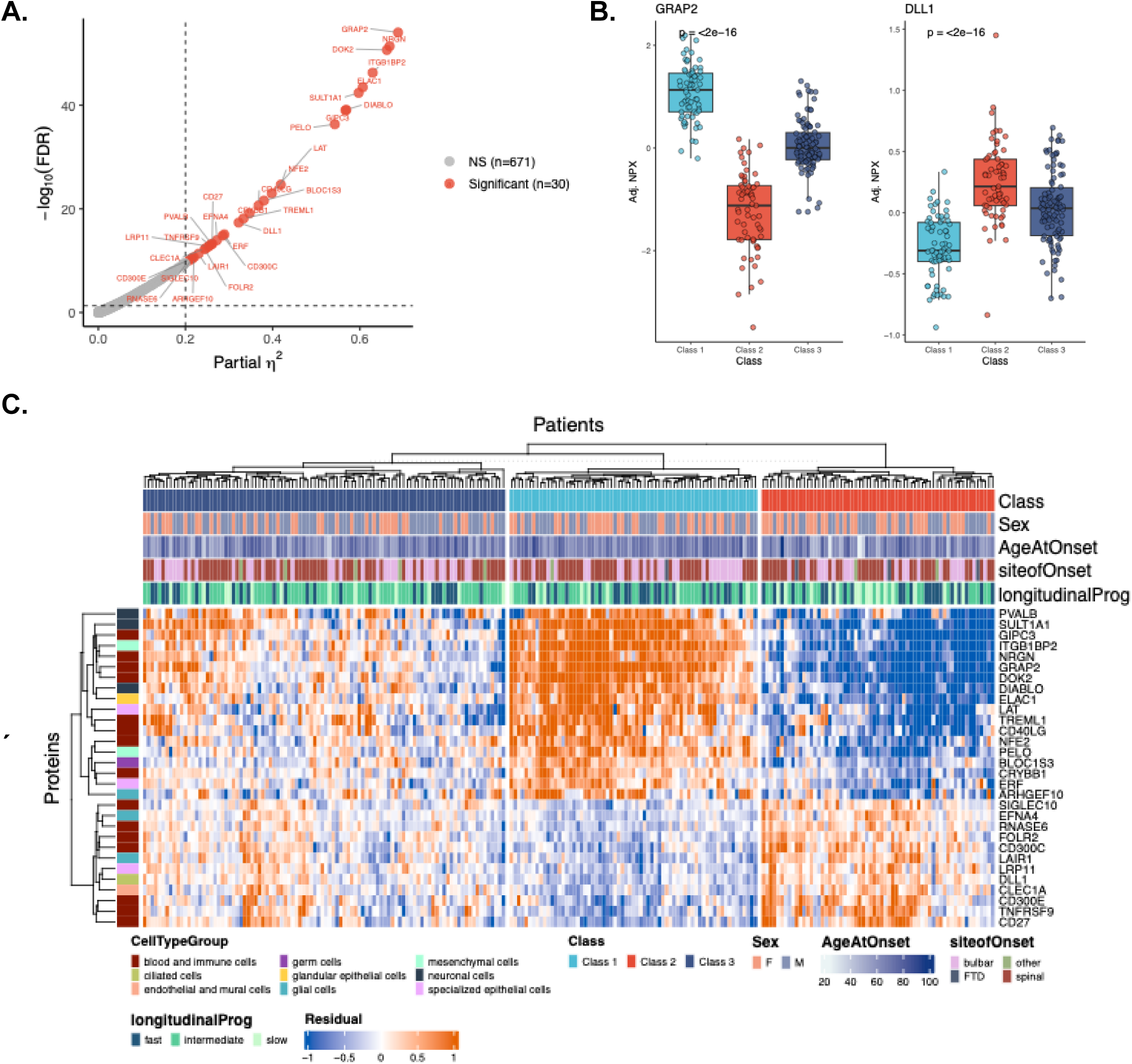
Cell-type markers associate with patient subgroups. **a.** Scatter plot showing the effect size (x-axis: partial η^2^) and −log_10_(FDR) values obtained from Type II ANOVA where each point represents a protein. Points colored in red represent proteins with FDR<0.05 (dashed horizontal line) and partial η^2^ > 0.20 (dashed vertical line), indicating that more than 20% of the variance in their levels is explained by patient subgroup membership. Points colored in grey (mentioned as NS) represent proteins that are non-significant considering the given thresholds. **b.** Boxplots showing the adjusted NPX values (y-axis) of two representative significant cell-type markers (GRAP2 representing Class 1 and DLL1 representing Class 2) across subgroups (x-axis) where each datapoint represents an individual. Boxes indicate interquartile range (IQR) with the median shown as the center line and whiskers represent 1.5 IQR. Boxplots and data points are colored in accordance with the patient subgroups. P-values were calculated by Type II ANOVA. **c.** Heatmap showing residual abundance of all significant cell-type enriched markers (rows) across patients (columns) and patient subgroups. Coloring of heatmap values correspond to high and low residual abundance, respectively. Annotation bars above the heatmap from top to bottom indicate patient subgroup (Class), sex, age at symptom onset (”ageatOnset”), site of symptom onset (“siteofOnset”) and disease progression category (“longitudinalProg”). The annotation bar (left) indicates the cell type group in which each protein is enriched, where enrichment is detected using datasets available on Human Protein Atlas (HPA) single-cell resource.

## Discussion

ALS is increasingly recognized as a biologically heterogenous disorder driven by multiple, partially overlapping pathogenic mechanisms^4,27–29^. While previous studies have defined molecular subgroups using postmortem transcriptomic profiling of motor- and frontal cortex, such approaches cannot be applied to living patients and do not capture circulating molecular signatures. Here, we profiled the plasma proteome across ALS patients with diverse site and age of onset, sex, and longitudinal progression. In alignment with previous studies, our plasma proteomic analysis identified three distinct molecular subgroups of ALS patients, each characterized by specific abundance patterns of protein modules representing diverse biological processes; inflammatory processes, transcriptional dysregulation, and cellular stress responses^17,19,21,30,31^.

Importantly, the abundance of these protein modules was associated with patient survival, demonstrating that circulating molecular profiles capture clinically meaningful aspects of disease progression. Moreover, we also linked plasma proteomic alterations to specific cell types by measuring overlapping markers between a single-cell RNA sequencing dataset and the Olink Explore HT Panel. Although this approach overlooks discrepancies between mRNA levels and protein abundance, we further showed that subgroup associated signatures may reflect contributions from nervous system cells, vascular cells and muscle cells, underscoring the multisystem nature of ALS.

Class 1 patients, characterized by elevated levels of turquoise, red and green modules, exhibited significantly shorter survival. Functional enrichment analysis indicated that these modules converge on biological processes that may contribute to the observed aggressive phenotype. Although plasma proteomics cannot determine tissue origin or confirm pathway activation, the enrichment of these pathways is consistent with mechanisms previously implicated in ALS. For instance, the turquoise module included proteins related to ERBB2 signaling, which is activated upon binding of NRG1 to ERBB3 or ERBB4, and participates in neuromuscular junction (NMJ) development and maintenance^32–35^. Earlier studies have also shown that NRG1 isoforms may reflect both glial activation and motor neuron damage^36,37^.Concurrent elevation of ITGB1BP, a muscle enriched protein, may suggest NMJ-related or muscle associated processes in Class 1 patients, although plasma levels alone cannot establish causality^38^. Interestingly, ERBB2 signaling further modulates the PI3K/AKT/mTOR pathway related red module, which controls global protein synthesis and is increasingly seen as a strong contributor to ALS pathogenesis^17,39,40^. Previous studies have also implicated Rho GTPases, including CDC42, in neurodegenerative processes including ALS^41^.

Beyond the signaling pathways, the hub protein of the turquoise module, ASCC1, forms the transcriptional co-regulator ASC-1 together with ASCC2, ASCC3 and TRIP4 to facilitate RNA processing as well as taking part in regulation of mRNA translation initiation downstream of mTOR signaling^42^. In previous studies, mutations of TRIP4 and ASCC1 have been shown to associate with spinal muscular atrophy (SMA) and neuromuscular function as well as congenital myopathy^43–45^. Despite this link, ASCC1 has never been identified as a plasma biomarker for ALS. Mouse studies have shown that *Ascc1* is expressed in dorsal root ganglia, paraspinal sympathetic ganglia and trigeminal ganglia^44^. Elevated plasma levels of ASCC1 accompanied by increased levels of PVALB, a marker of proprioceptive neurons found in dorsal root ganglia, where they mediate sensorimotor communication between muscle and the central nervous system, raise the possibility that peripheral sensory circuits may contribute to the circulating proteomic signature in Class 1^46–48^. Although intriguing, validation using post-mortem tissue or longitudinal sampling is needed to explain if the observed alterations reflect neuronal degeneration, altered secretion or peripheral tissue involvement.

In contrast to Class 1 patients with shorter survival outcomes, Class 2 and Class 3 patients exhibited significantly longer survival when compared to Class 1. While Class 3 patients had survival outcomes similar to Class 2 patients, we observed that Class 3 patients did not have any significant correlation with the identified protein modules or the cell-type enriched markers. Class 2 patients exhibited elevated levels of the pink, black, brown and blue modules and had significantly longer survival compared to Class 1. These modules were enriched for biological processes related to synaptic transmission, vascular function immune signaling, and cell maturation. Elevated levels of PTPRR, the hub protein of the pink module, have been detected in postmortem cortex samples and associated with psychiatric phenotypes, although, its role in neurodegeneration remains unclear^49^. The pink module also included GFAP, an astrocyte marker increasingly recognized as a plasma biomarker of neurodegenerative diseases, importantly dementias^50–52^. Despite the association of GFAP with neuroinflammation Class 2 patients exhibited longer survival than Class 1, suggesting that inflammatory markers may not directly mirror central nervous system specific pathology and may instead reflect systemic immune states or vascular processes^52^. Nevertheless, concurrent elevation of pink and black modules as well as vascular endothelial cell markers such as CLEC1A, a part of blue module, may indicate blood-brain barrier involvement or peripheral endothelial activation in Class 2 patients. An interesting future direction would be to test whether elevated GFAP and PTPRR signatures in Class 2 patients correlate with cognitive dysfunction.

Inflammation is increasingly seen as a prominent mechanism driving ALS heterogeneity. TGFBR2 emerged as a hub protein of the brown module, which was highly abundant in Class 2 patients. A previous study has implicated TGF-ß signaling in inflammatory processes associated with astrocytes and ALS^53^. Combined with increased levels of GFAP, the elevation of TGFBR2 may suggest that the inflammatory signature of Class 2 is driven by astrocyte related processes. GO analysis further identified enriched terms including cell-cell adhesion, transmembrane-ephrin receptor activity, and the TNFR2 non-canonical NF-κB pathway for the brown module. Activation of non-canonical NF-κB signaling by TNF superfamily members has been linked to neuroinflammation in the central nervous system^54^. Unlike Class 1 patients, Class 2 patients had elevated levels of microglia markers EFNA4 and LAIR1. Interestingly, Class 1 was characterized by elevated adaptive immune cell markers, while Class 2 showed elevated innate immune cell markers. One interpretation could suggest that Class 2 and Class 1 reflect different biological trajectories, where Class 2 represents a state characterized by active innate immune responses and Class 1 by more advanced or distinct stage with adaptive immune response involvement. However, similar ALSFRS-R values between Class 1 and Class 2 support the interpretation that these subgroups represent fundamentally distinct disease mechanisms rather than disease stages as also evidenced by the time of survival after symptom onset where we observe faster deterioration in Class 1 patients. These findings suggest a need for further investigations to understand the interplay between innate and adaptive immune responses both in peripheral tissues and the central nervous system across ALS subgroups more broadly.

While plasma proteomics cannot determine tissue origin or casual mechanisms and the design of this study limits the ability to assess temporal changes, our results demonstrate that ALS can be stratified into biologically distinct subgroups defined by plasma proteomic signatures, supporting the concept of ALS as a molecularly heterogeneous disease. Importantly, these proteomic subgroups and their associated protein modules are linked to survival, indicating that circulating molecular profiles capture clinically meaningful aspects of disease progression. Altogether, these findings represent potential implications for patient stratification in clinical trials, prognostic biomarker development, and mechanistic discovery.

## Data Availability

Dataset used in this study is available from senior authors upon reasonable request.

## Code Availability

All code and scripts are available on Zenodo (xxxxxx) and Månberg Lab’s GitHub repository (xxxxxx).

## Acknowledgements

We would like to thank all patients and the families for their invaluable contribution to this work. We also thank all team members, especially Jenny Hellqvist, for their efforts in sample and data collection.

## Methods

### Study design and ethics

This study was based on cross-sectional samples collected from ALS patients from the Stockholm Region who were part of the ALSrisc Study^22^. The study was conducted in accordance with the Declaration of Helsinki. Written informed consent to participate the study was obtained from all individuals and the study was reviewed and approved by the Swedish Ethical Review Authority (DNRs. 2017/1895-31 and 2018/1605-31).

### Cohort description and sample collection

Included patients were newly diagnosed with ALS according to the El Escorial Criteria or Gold Coast Criteria between February 2016 - September 2023 and had at least three amyotrophic lateral sclerosis rating scale revised (ALSFRS-R) measurements (**Table 1**)^55^. The clinical information collected included site, date and age of symptom onset, biological sex, body mass index (BMI) and ALSFRS-R scores. ALSFRS-R is a validated 12-item scale ranging between 48 and 0 that assesses functional severity and where maximum value represents normal function^56^. ALSFRS-R was collected every six months for each patient and progression rate was calculated as follows:

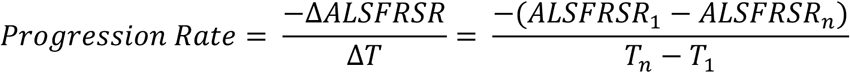

Where Δ*ALSFRSR* represents the change of score between the first and last ALSFRS-R scores assessed at Karolinska University Hospital and Δ*T* represents the time between last and first assessments as months. The patients were divided into three categories based on their progression rates as slow (*Progression Rate* < 0.5 *score*/*mont*ℎ), intermediate(0.5 *score*/*mont*ℎ ≤ *Progression Rate* < 1.5 *score*/*mont*ℎ) and fast (*Progression Rate* ≥ 1.5*score*/*mont*ℎ) progressors. Survival endpoints were defined as death or the date of tracheostomy; patients who were alive and free of tracheostomy at the last follow-up in October 2025 were censored.

Baseline plasma samples from the included individuals were collected by phlebotomy using EDTA tubes within a median of 36 days from the date of diagnosis followed by centrifugation and removal of blood cells. The samples were stored at −80°C before analysis.

### Plasma proteome profiling using Olink Explore HT Assay

The Olink Explore HT assay is a recently developed Proximity Extension Assay (PEA) platform that allows the simultaneous, high-throughput measurement of approximately 5400 proteins. The technology utilizes the oligonucleotide coupled, paired antibodies which bind to analytes abundant in various specimens including, but not limited to, plasma and serum. When the antibodies are in close vicinity, the oligonucleotide chains form a double-stranded DNA chain which is amplified and quantified by Illumina NovaSeq. The plasma proteome from each individual was measured using Olink Explore HT Assay at SciLifeLab Affinity Proteomics and National Genomics Infrastructure (NGI) in Uppsala, Sweden, as part of the Human Disease Blood Atlas (HDBA) project^8^.

Initial quality control included assessments of incubation, extension, detection and interplate reference samples. Assay measurements that did not pass the predefined thresholds were flagged. Following the quality control, data was normalized in accordance with the intensity normalization procedure, minimizing the technical variation across samples and plates. Normalized Protein Expression (NPX) values, which represent relative protein abundance and enables comparison of the same protein across samples, were generated using the NPX map software by using internal control samples to adjust for technical variation across samples and plates.

Furthermore, all data points failing quality control were removed prior to downstream analysis. Samples with >50% of measurements flagged for quality control (QC) failure were excluded. Proteins from dilution block 8 (n = 68, dilution 1:100,000) were excluded due to technical issues identified by the provider.

Assays that were above limit of detection (LOD) in at least 85% of samples were included in the study. Principal component analysis (PCA) was performed using the “pca” function of PCAtools (v.2.18.0) R/Bioconductor Package (parameters: removeVar =0.25, scale=T) on NPX values of included assays^57^. Elbow point was determined using “findElbowPoint” function and screeplot was generated using “screeplot” function with the determined elbow point and percent explained variation by each of the principal components assessed. PCA plots were generated using “biplot” function from the same package. The relationship between the top 10 principal components (PC1-10) and technical/clinical covariates were assessed and plotted using “eigencorplot” function and significance was determined using False Discovery Rate (FDR) (parameters: components = getComponents(pcaObj, 1:10, scale =TRUE, plotRsquared =TRUE, corFun = “pearson”, corUSE = “pairwise.complete.obs”,corMultipleTestCorrection =”BH”,posLab=”all”).

NPX values for each protein were adjusted for identified confounders (median NPX value, age and biological sex) using multivariate linear regression:

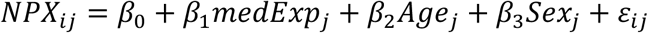

Where *NPX_i_*_j_ denotes the NPX value of protein *i* in individual *j*. Per individual and per protein residuals from each model were used as covariate-adjusted NPX values in subsequent analyses.

### Consensus clustering

To classify patients based on plasma proteome, adjusted NPX values were obtained from the multivariate linear regression models and used as input in the cola (v.2.12.0) R/Bioconductor package^58^. “Adjust_matrix” function was used to remove assays with low variance, and “run_all_consensus_partition_methods” function was used to identify optimal clustering solutions (parameters: max_k = 10, top_n=nrow(matrix)*0.25). Solutions with k > 2 were further assessed based on 1-PAC, mean silhouette and concordance values, suggesting the combination of ability to correlate to other rows (ATC) variable selection and kmeans clustering as the most optimal clustering solution based on the determined criteria (**Suppl. Fig. 2B**). Following this, clustering was performed using “consensus_partition” function (set.seed(121) and parameters: top_value_method: “ATC”, partition_method=”kmeans”, max_k = 10, scale_rows=TRUE, top_n = nrow(matrix)*0.25). Proteins that had different levels across identified patient subgroups were extracted using “get_signatures” function (parameters: fdr_cutoff=0.05, silhouette_cutoff = 0.7, group_diff = 0.25) (**Suppl. Fig. 2F**). Following this, a heatmap showing identified proteins (n = 1466) and patients with silhouette value > 0.7 (n=228) was generated using “Heatmap” function from Com-plexHeatmap (v. 2.22.0) R/Bioconductor package and a UMAP plot colored by patient classes was generated using “umap” (set.seed(123) and parameters: n_neighbors = 15, min_dist = 0.1) function from umap (v. 2.10.0) R package^59,60^.

Survival of the identified patient subgroups was assessed by Kaplan-Meier estimates using “survfit” function from survival (v.3.8-3) R package, with time-to-event measured in months from the date of symptom onset^61^. Survival curves including 95% confidence intervals and risk tables were visualized, and survival outcomes of subgroups were compared by log-rank test using “ggsurvplot” function (parameters: pval = TRUE, risk.table = TRUE,conf.int = TRUE) from survminer (v.0.5.0) R package^62^.

### Weighted Gene Correlation Network Analysis (WGCNA)

Weighted Gene Correlation Network Analysis (WGCNA) is a computational, network-based approach to identify modules of genes or proteins that exhibit co-expression patterns across samples. WGCNA was performed using adjusted NPX values of 1466 proteins across 228 patients to identify protein modules with distinct abundance patterns in plasma across patient subgroups using functions from WGCNA (v. 1.73) R/Bioconductor package^63^. Quality of the dataset for WGCNA analysis was assessed using “goodSamplesGenes” function with default parameters. Following this, scale-free topology was assessed across a range (1 to 20) of soft-thresholding powers and 4 was estimated as the appropriate soft-thresholding power using “pickSoftThreshold” function (**Suppl. Fig. 4A**). “blockwiseModules” function was used with default parameters to construct networks and detect modules. Dendrogram of modules was plotted using “plotDendroAndColors” function with default parameters (**Suppl. Fig. 4B**). Top hubs for each of the detected modules were identified using “chooseTopHubInEachModule” function with default parameters. Module eigengenes (MEs), representing the first principal component of each module across patients, were further used for module-trait association between protein modules and the patient subgroups. Module-trait association r values were calculated using “cor” function (parameters: method=”pearson”) and the significance of correlations were assessed using “corPValueStudent” function with default parameters. The correlation heatmap was visualized using “Heatmap” function from ComplexHeatmap R/Bioconductor package.

To assess whether the protein modules correlate with survival outcome, optimal cut-point for ME values were determined using maximally selected rank statistics implemented by “surv_cutpoint” (parameters: minprop: 0.15) function of survminer (v. 0.5.0) R package with time-to-event measured in months from date of symptom onset. “surv_categorize” function of survminer R package was then used to categorize patients into “high” and “low” groups for a given module based on the determined cut-point. Survival outcome of patients in “high” and “low” groups was assessed by Kaplan-Meier estimates using “survfit” function from survival R package, with time-to-event measured in months from the date of symptom onset. Survival curves including 95% confidence intervals and risk tables were visualized, and survival outcomes of subgroups were compared by log-rank test using “ggsurvplot” function (parameters: pval = TRUE, risk.table = TRUE,conf.int = TRUE) from survminer R package.

### Functional Enrichment Analysis

To relate modules to known biological functions, right-sided hypergeometric test was performed using Cytoscape (v.3.10.3) plug-in ClueGO (v.2.5.10) together with p value cutoff = T, Correction Method Used = Benjamini-Hochberg, midPValues = T, Statistical Test Used = Enrichment (Right-sided hypergeometric test), Kappa = 0.4, Min.Percentage = 4%, Min#Genes = 3, GO Fusion = TRUE, Initial Group Size = 1, Sharing Group Percentage = 50, Leading Group Term based on = Highest Significance, GO Tree Interval Min Level=3, Max Level= 8 Ontology Used = GO_BiologicalProcess-EBI-Uni-Prot-GOA-ACAP-ARAP_25.05.2022 & GO_MolecularFunction-EBI-UniProt-GOA-ACAP-ARAP_25.05.2022, REACTOME_Pathways_25.05.2022 with a Custom Reference Set consisted of the 1466 proteins that were used for WGCNA analysis^64^. FDR threshold of 0.05 was used for all modules except for the blue (FDR < 0.10) and the turquoise (FDR <0.01) modules.

### Cell-type enrichment analysis

The Human Protein Atlas (HPA) is an initiative which uses various omics data to map human proteins in cells, tissues and organs^65^. The single-cell resource within the initiative provides vast information about gene expression in 154 cell types^66^. HPA v.25 which provides normalized counts per million (nCPM) for each transcript in single-cells was used to infer relative abundance of cell-type specific markers in plasma and whether these markers could differentiate between patient subgroups. First, the transcripts expressed in fewer than 25% of cell types were removed to limit the analysis to the transcripts detectable across cell types. Tau-specificity score is a quantitative measure ranging between 0 and 1 indicating the specificity of a transcript or protein across cells and tissues where values closer to 1 indicate high specificity^67^. For each transcript that passed the filtering criteria, tau-specificity score was calculated using the log-transformed nCPM values as described by Yanai et al. and HPA single-cell resource. Markers with tau scores larger than 0.85 were classified as cell-type enriched. Following this, the transcripts were mapped to UniProt identifiers and were then restricted to proteins measured by the Olink Explore HT assay.

To assess whether the selected markers are enriched in patient subgroups, a linear model was fitted for each marker across patients:

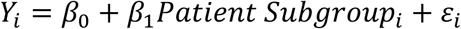

Where *Y_i_* denotes the adjusted NPX value in patient *i* and *Patient Subgroup_i_* denotes the group membership of patient *i*. Type II Analysis of Variance (ANOVA) was performed using “anova” function from car (v.3.1.3) R package^68^. The effect sizes (h^2^) which represent the variance in adjusted NPX values explained by patient subgroups were calculated using “eta_squared” function from effectsize (v.1.0.0) R package^69^. Statistical significance was assessed by p-values obtained from the ANOVA which were further adjusted for multiple testing using false discovery rate (FDR) method using “p.adjust” function from stats (v.4.4.1) R package. Markers with FDR < 0.05 and partial h^2^ > 0.20 were considered to alter significantly between patient groups and linked to biological context and cell-types using information readily available on Human Protein Atlas resource (**Suppl. Fig. 8**)^64^. Protein-level plots were generated using ggplot2 R package (v.3.5.2) and heatmaps showing marker levels in individual patients were generated using ComplexHeatmap R package^59,70^.

---

**Supplementary Figure 1:**
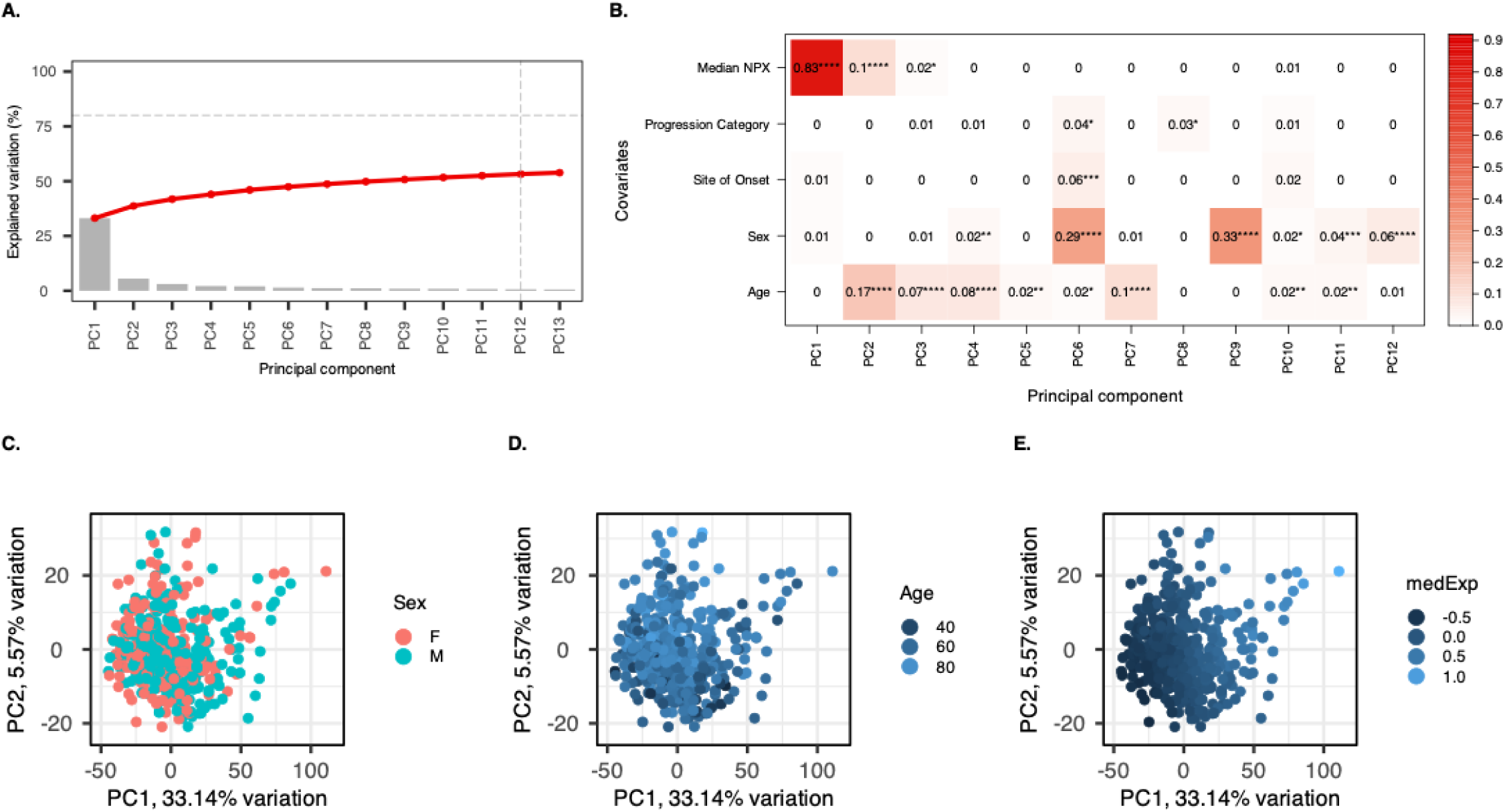
Principal component analysis of plasma proteome indicates technical and biological variation. **a.** Scree plot showing the variance explained (y-axis) by each principal component (PC) represented in grey colored bars along x-axis. Red line represents the cumulative sum of variance explained by all PCs. Horizontal line represents cumulative variance threshold (80%) whereas the vertical line represents the PCs that explain highest cumulative amount of variation in the data. **b.** Eigen-correlation plot showing the correlation between PC eigen values (columns) and technical and clinical covariates (rows). Values written in the cells represent Pearson correlation coefficients while coloring of cells represents the magnitude of absolute associations between the PCs and the covariates. Asterisks denote significance of correlations (“****” ∼ 0, “***” < 0.0001, “**” < 0.01, “*” < 0.05, none > 0.5). **c, d, and e.** Biplots showing PC1 (x-axis; explains 33.14% of variation) and PC2 (y-axis; explains 5.57% of variation) colored by sex, age and median NPX value, respectively.

**Supplementary Figure 2:**
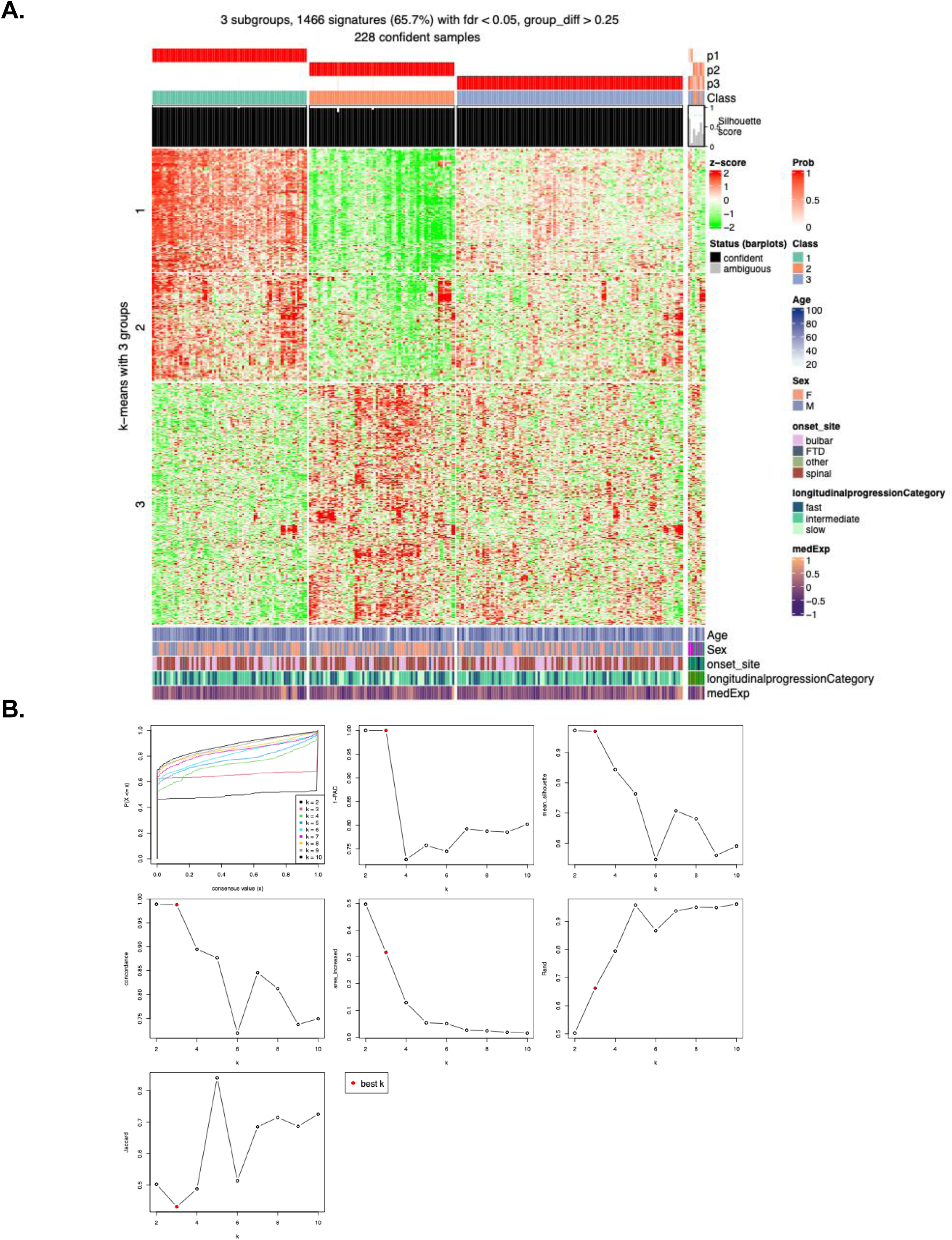
“cola” clustering determines the optimal clustering solution. **a.** Heatmap showing patient clustering using cola – ATC, kmeans combination with k=3. Columns represent patients (n=228) split by patient subgroups whereas rows represent 1466 proteins differentially abundant proteins (FDR < 0.05, F-test and |log2FC| > 0. 25) across the patient groups divided into three sets. Horizontal red bars at the top (p1, p2, p3) represent the probability of each patient being assigned to Class 1, 2 or 3 across subsampling iterations. Vertical black and grey bars represent the silhouette scores for patients that are confidently (n=228, silhouette score > 0.7) and ambiguously (n=7, silhouette score < 0.7) clustered, respectively. Horizontal annotation bars below the heatmap indicate age at symptom onset, sex, site of symptom onset (“onset_site”), disease progression category (“longitudinalprogressionCategory”), and median NPX value(”medExp”) for each patient. **b.** Consensus clustering statistics across k = 2-10 indicating stable clustering at k=3. Panels show the empirical cumulative distribution function (ECDF) of consensus values, 1-PAC (proportion of ambiguous clustering), mean silhouette width, concordance, area under ECDF increase. Rand index, and Jaccard index. Dots colored red indicate the optimal k.

**Supplementary Figure 3:**
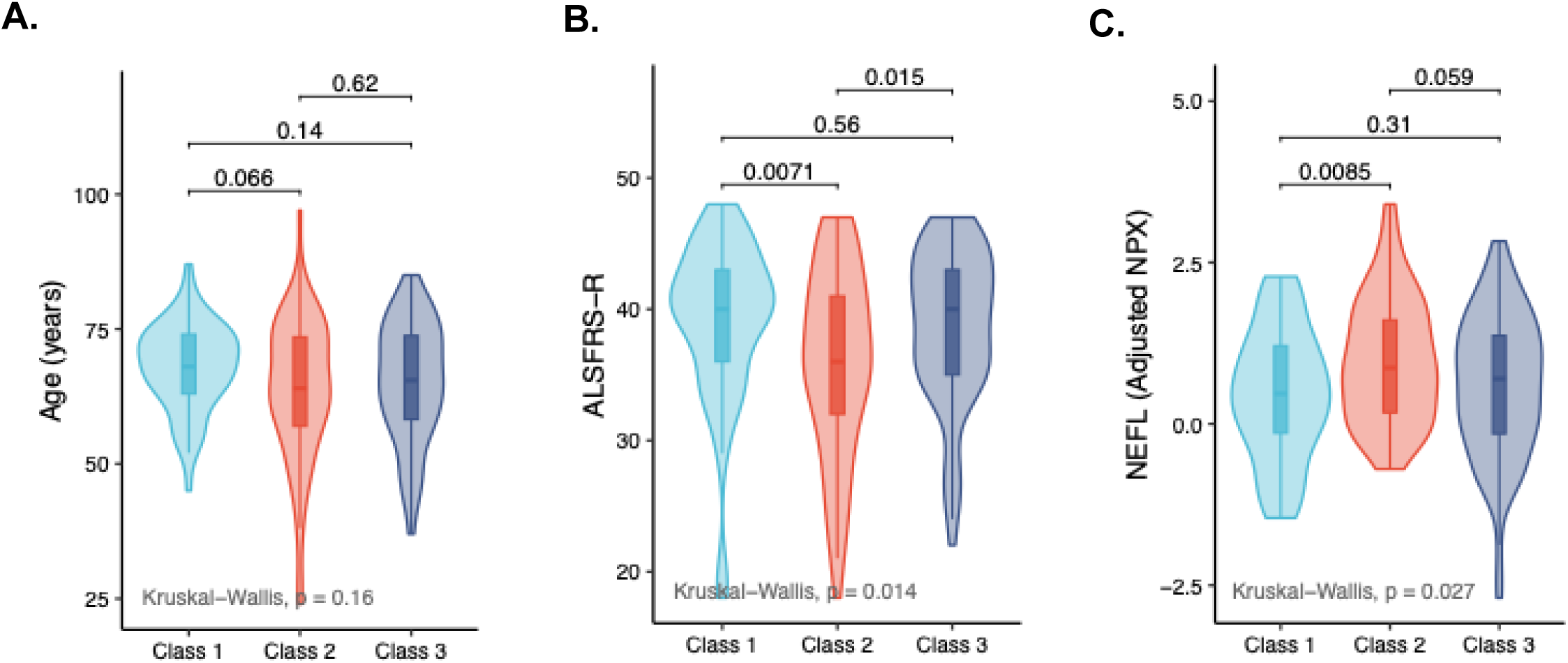
ALSFRS-R scores, Age and adjusted NPX values for NEFL across patient subgroups. Violin plots showing the distribution of **a.** Age (years; y-axis), **b.** ALSFRS-R score (y-axis) at first visit, and **c.** NEFL (Adjusted NPX; y-axis) across Class 1, Class 2 and Class 3 patients (x-axis). The width of each violin represents the density of data points at that value. Internal boxes colored in accordance with the patient subgroups indicate interquartile range (IQR) with the median shown as the center line and whiskers represent 1.5 IQR. Pairwise comparisons were performed using the Wilcoxon rank sum test while overall group differences were assessed using Kruskal-Wallis test (Age: p = 0.16; ALSFRS-R: p = 0.014 and NEFL: p= 0.027).

**Supplementary Figure 4:**
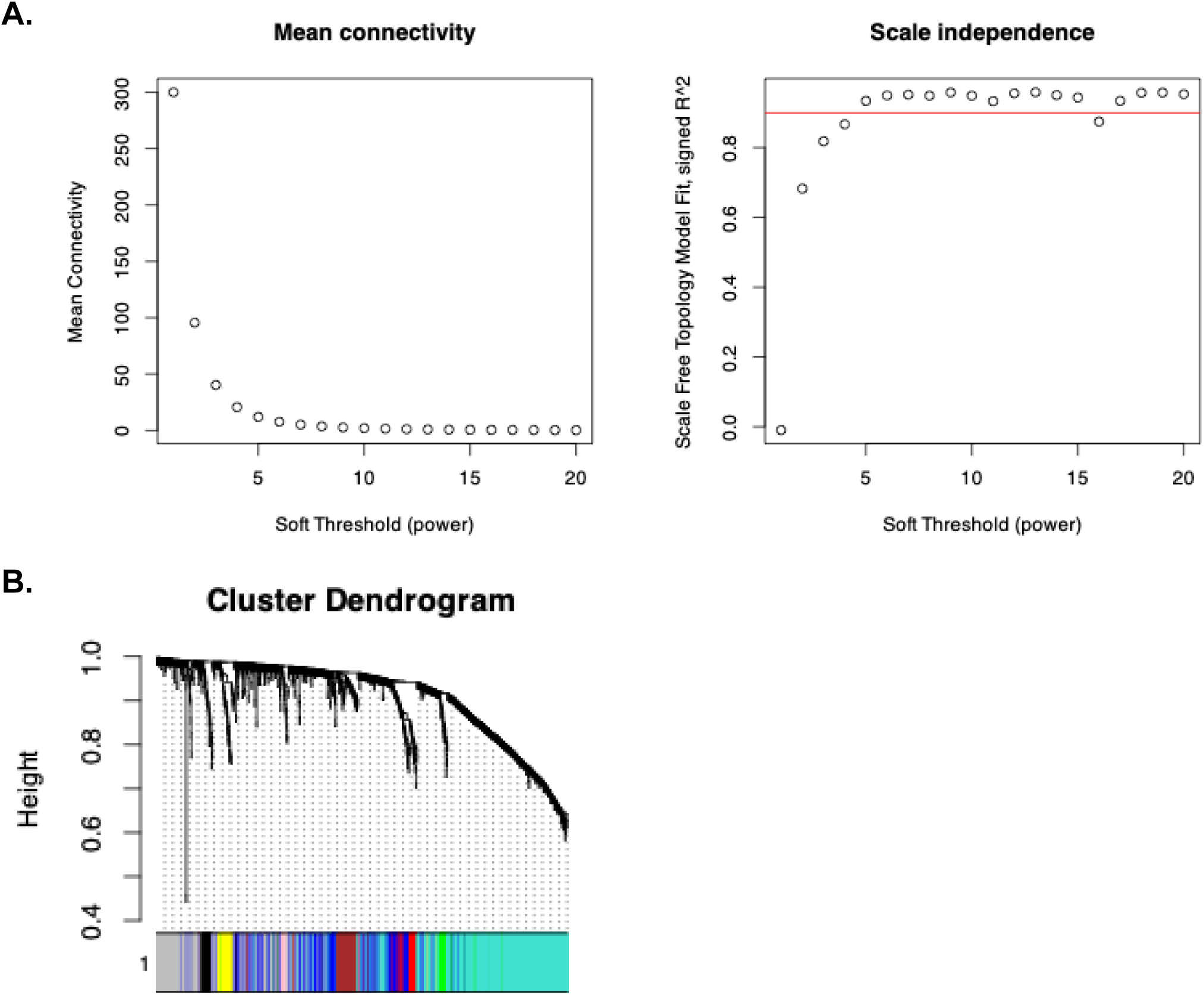
Network parameters for WGCNA. **a.** Scatter plot showing scale free topology fit index (R^2^;y-axis) and soft thresholding power (x-axis) where scale-free topology fit index serves as a measure of how well the network follows a scale-free pattern. The red horizontal line represents the R^2^ threshold used for choosing the soft-thresholding power. **b.** Cluster dendrogram of the 1466 proteins that significantly differ across ALS subgroups constructed using a topological dissimilarity matrix with soft-thresholding power of 4. The colored horizontal bar below the dendrogram indicates module assignments of individual proteins.

**Supplementary Figure 5:**
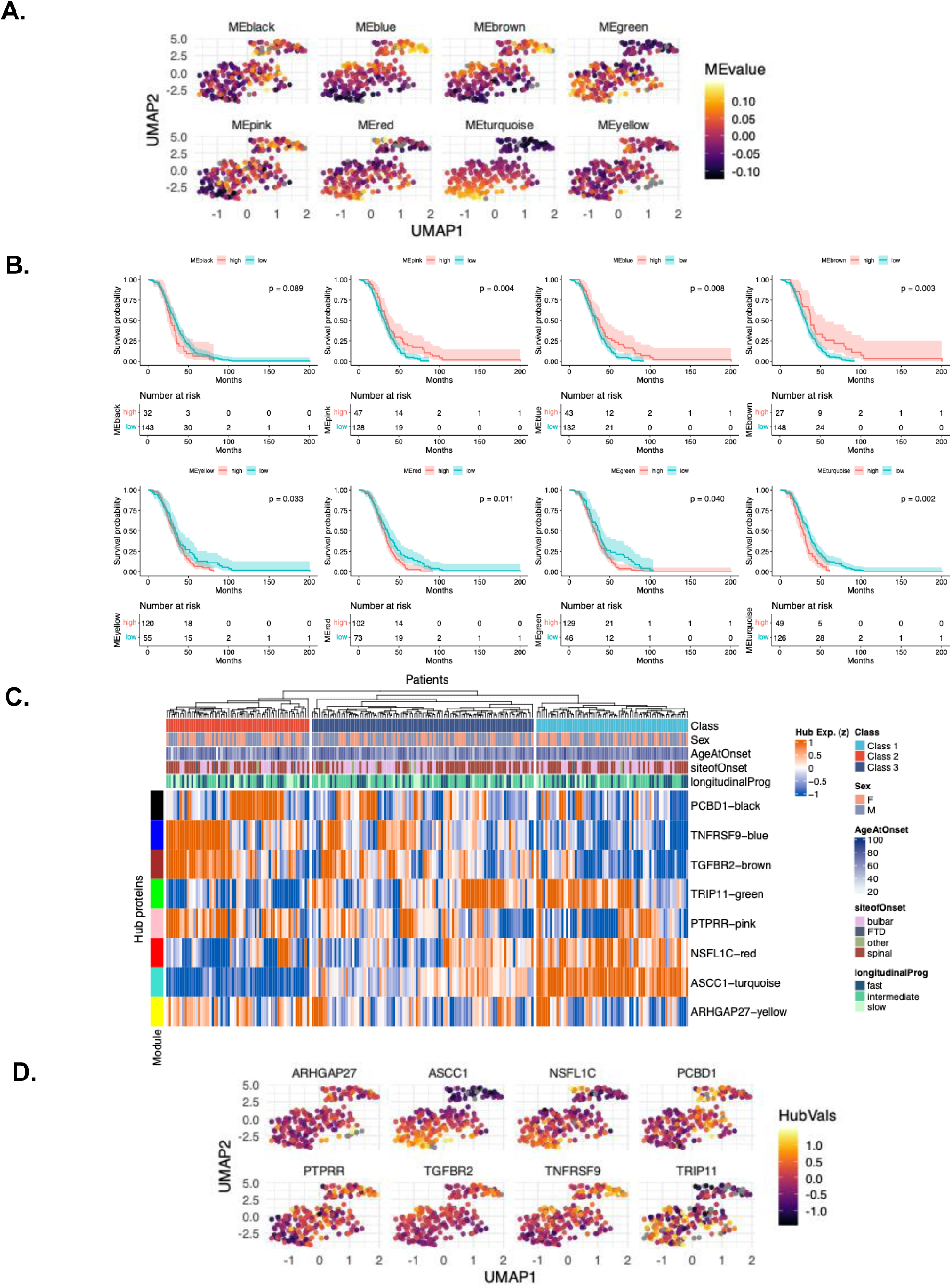
Module abundances across patients. **a.** UMAP of ALS patients (n=228) generated using adjusted abundance values of significantly altered plasma proteins, where each dot represents a patient colored by the module eigengene (ME) value of each WGCNA module. For each module higher ME value is indicated by yellow/orange color while lower ME value is indicated by dark purple color. **b.** Kaplan-Meier survival curves stratified into “high” (in red) and “low” (in teal) groups by ME values based on optimal cutoffs shown for all eight modules. The p-values were calculated using Log-rank test, error bands represent 95% confidence interval and tables below each panel represent number of individuals at risk for “high” and “low” groups. **c.** Heatmap showing residual abundance of all significant markers across patients (columns) and patient subgroups (top annotation). Coloring of the heatmap represents the residual abundance of each module hub (rows) across patients, red indicates high abundance whereas blue indicates low. Annotation bars above the heatmap from top to bottom indicate patient subgroup (Class), sex, age at symptom onset (”ageatOnset”), site of symptom onset (“siteofOnset”) and disease progression category (“longitudinalProg”). **d.** UMAP of ALS patients (n=228) generated using adjusted abundance values of significantly altered plasma proteins, where each dot represents a patient colored by the residual abundance of each WGCNA module hub. For each hub protein, higher residual abundance is indicated by yellow/orange color while lower residual abundance is indicated by dark purple color.

**Supplementary Figure 6:**
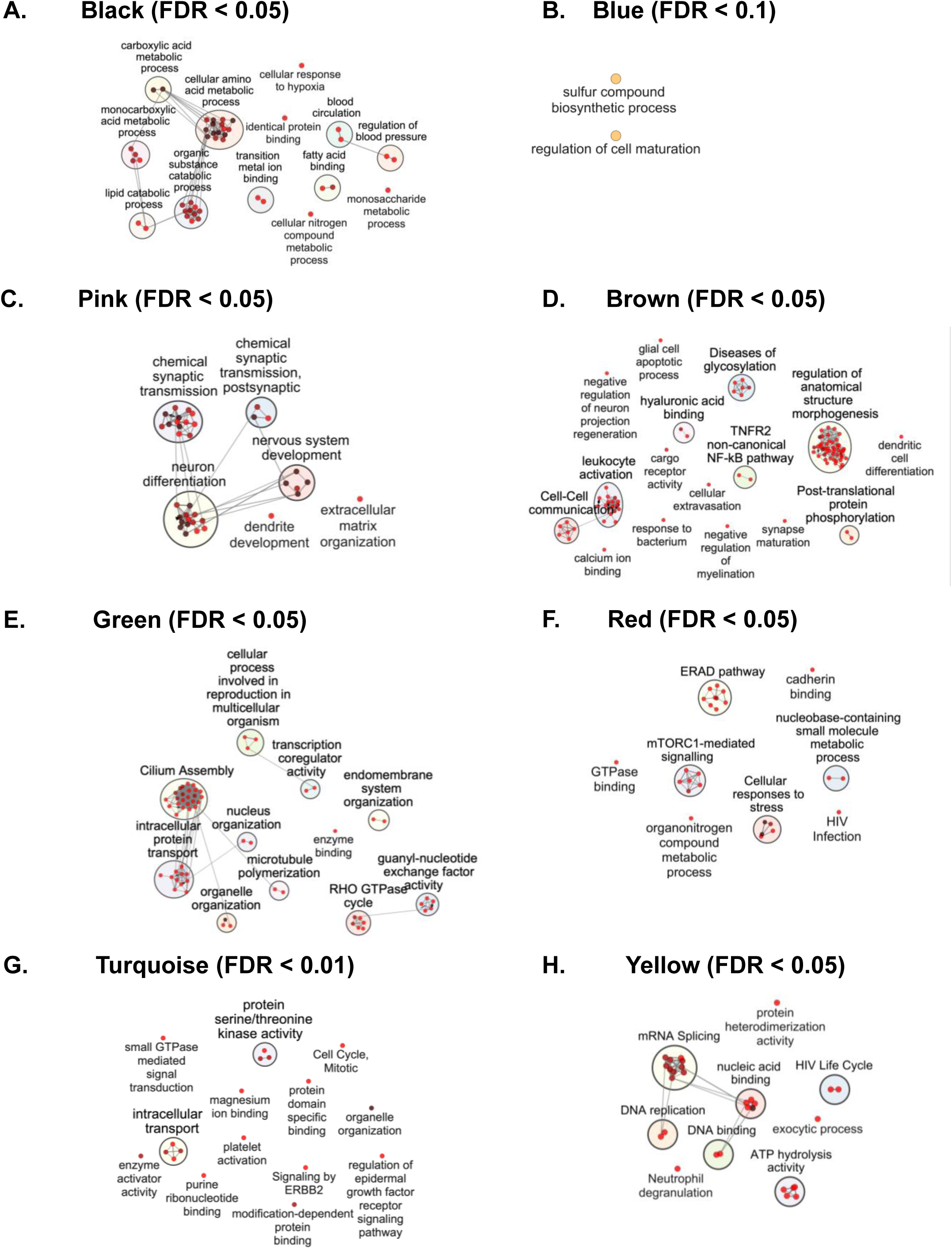
Networks of biological functions associated with patient subgroups. Networks of enriched biological processes and pathways associated with each protein module generated using ClueGO. Enrichment was assessed using a right-sided hypergeometric test with Benjamini-Hochberg correction for multiple testing. Each panel is labeled with the corresponding module name, and the applied FDR threshold is indicated in parenthesis (**a**. Black, FDR <0.05; **b**. Blue, FDR < 0.1; **c**. Pink, FDR< 0.05; **d**. Brown, FDR < 0.05; **e**. Green, FDR < 0.05; **f**. Red, FDR < 0.05; **g**. Turquoise, FDR < 0.01 and **h**.Yellow, FDR < 0.05). For each network, node represents a biological process or pathway, and edges connect functionally related terms within the same cluster. Node size reflects the number of proteins associated with each term, and node color intensity indicates the false discovery rate (FDR). Functionally related terms are grouped into clusters, encircled together, and the most statistically significant term within each cluster is used as the cluster label.

**Supplementary Figure 7:**
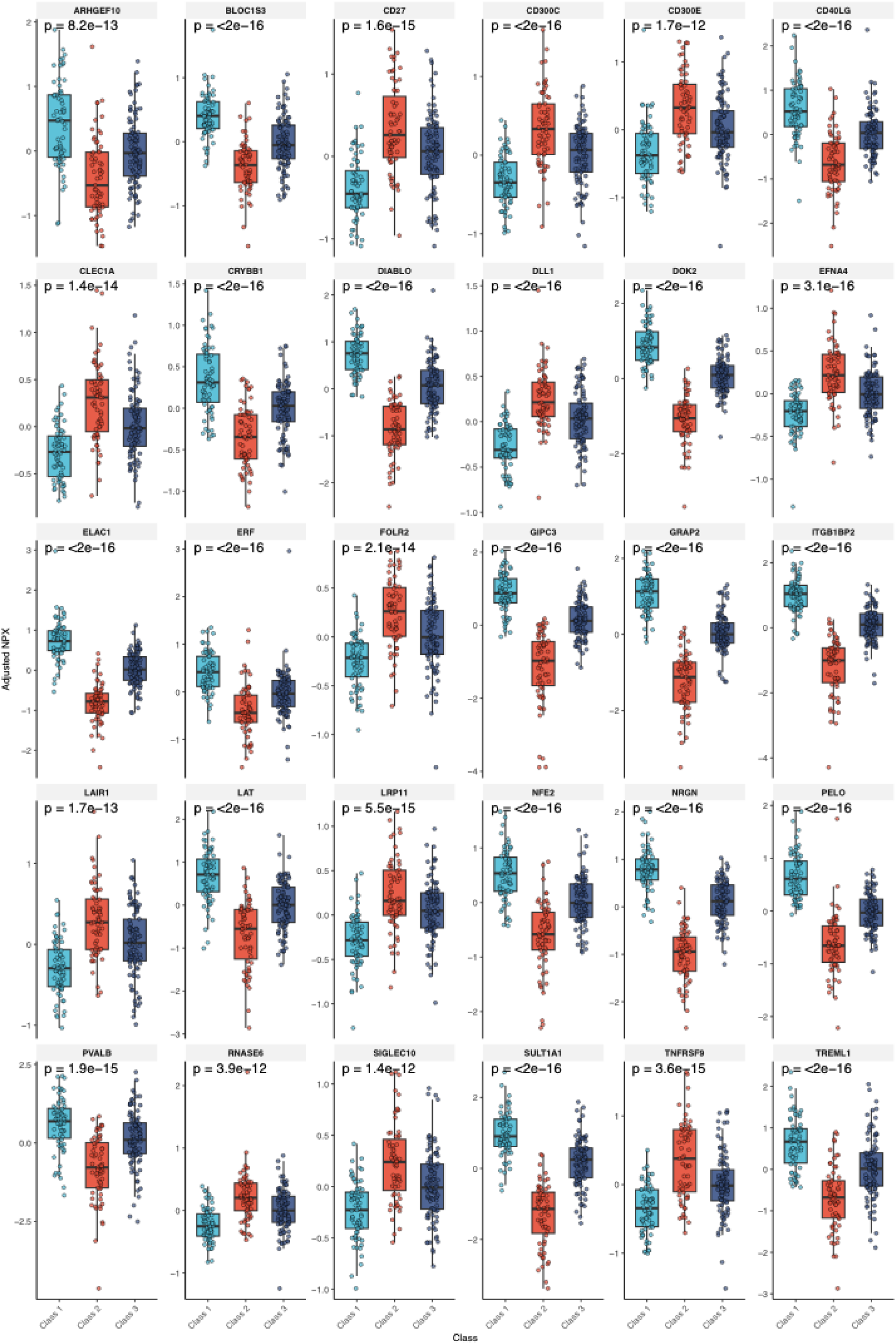
Cell-type enriched markers across patient subgroups. Boxplots showing the adjusted NPX values (y-axis) of significant cell-type markers (FDR < 0.05, Type II ANOVA and and partial η^2^ > 0.20) across subgroups (x-axis) where each datapoint represents an individual. Boxes indicate interquartile range (IQR) with the median shown as the center line and whiskers represent 1.5 IQR. Boxplots and data points are colored in accordance with the patient subgroups. P-values were calculated by Type II ANOVA.

**Supplementary Figure 8:**
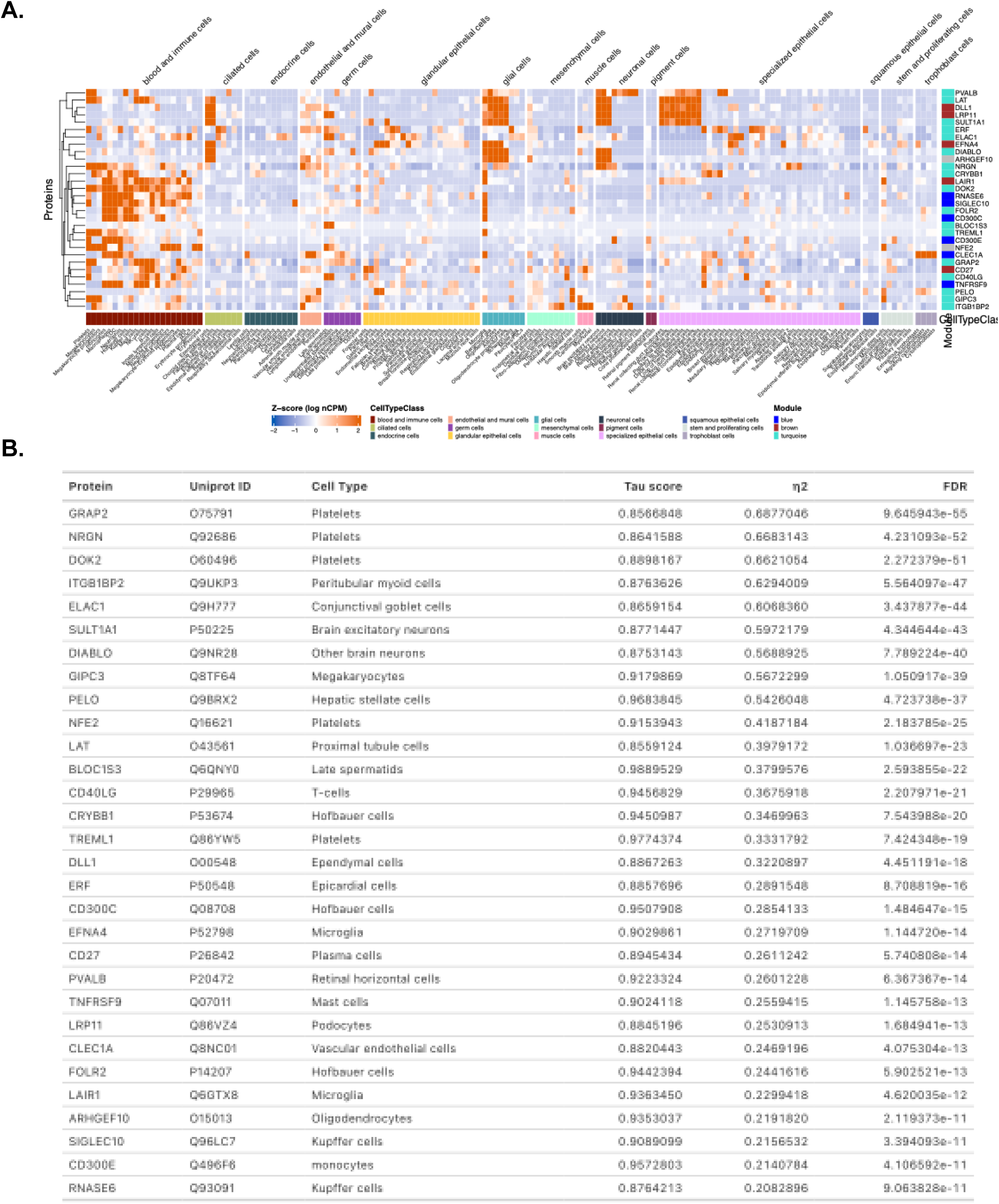
Functional context of cell-type enriched markers that can discriminate between patient subgroups. **a.** Heatmap of z-scored nCPM values obtained from the Human Protein Atlas (HPA) for 30 cell-type associated markers across 154 cell types grouped by the cell type class (column annotation; under the heatmap). Markers (rows) are hierarchically clustered and colored by WGCNA module membership (row annotation; right). **b.** Table of cell-type associated markers that have altered levels across subgroups. Partial eta squared (η^2^), effect size, corresponds to variance in adjusted NPX values of cell-type associated markers that can be explained by patient groups. Tau score indicates specificity the marker for a given cell-type. P-values were calculated using Type II Anova and were FDR-adjusted.

